## Supplementary material for "Digital delivery of Behavioural Activation therapy to overcome depression and facilitate social and economic transitions of adolescents in South Africa (the DoBAt study): protocol for a pilot randomised controlled trial": DoBAt Study Supplementary Materials

### Table 1. Investigators' affiliations and contact details

| Name | Role | Affiliation | Email |
|---|---|---|---|
| **Prof. Kathleen Kahn** | Co-Principal Investigator | Professor of Public Health and Principal Scientist, MRC/Wits Rural Public Health and Heath Transitions Research Unit (Agincourt), School of Public Health, University of the Witwatersrand, Johannesburg, South Africa | |
| **Prof. Alan Stein** | Co-Principal Investigator | Professor and Head of Section, Child and Adolescent Psychiatry, Department of Child and Adolescent Psychiatry, University of Oxford, Oxford, United Kingdom | |
| **Prof. Sarah-Jayne Blakemore** | Investigator | Professor of Psychology and Cognitive Neuroscience, University of Cambridge, Cambridge, United Kingdom | |
| **Dr. Gabriele Chierchia** | Investigator | Research and Teaching Associate at the University of Cambridge Department of Psychology, Cambridge, United Kingdom | |
| **Prof. Michelle Craske** | Investigator | Professor of Psychology, Psychiatry and Biobehavioral Sciences at the University of California, Los Angeles (UCLA), United States of America | |
| **Ms Sophie Luise Fielmann** | Investigator | Doctoral Student, Department of Psychology, University of Cambridge, Cambridge, United Kingdom | |
| **Dr. Xavier Gómez-Olivé** | Investigator/ Agincourt Research Manager | Research Manager, MRC/Wits Rural Public Health and Heath Transitions Research Unit (Agincourt), School of Public Health, University of the Witwatersrand, Johannesburg, South Africa | |
| **Prof. Alastair van Heerden** | Investigator | Associate Professor University of Witwatersrand, and Research Director at Human Sciences Research Council (HSRC), Pietermaritzburg, South Africa | |
| **Dr. Emma Kilford** | Investigator | Postdoctoral research fellow at University College London (UCL) Department of Clinical, Educational and Health Psychology, London, United Kingdom | |
| **Prof. Crick Lund** | Investigator | Professor of Global Mental Health and Development, Centre for Global Mental Health, Institute of Psychiatry, Psychology and Neuroscience, King's College London, United Kingdom; Honorary Professor of | |

| | | | |
|---|---|---|---|
| | | Public Mental Health, Alan J Flisher Centre for Public Mental Health, Department of Psychiatry and Mental Health, University of Cape Town, South Africa | |
| **Prof. Heather O'Mahen** | Investigator | Associate Professor in Perinatal Clinical Psychology, University of Exeter, Exeter, United Kingdom | |
| **Mr Daniel Mahlangu** | Investigator | Data Specialist DoBAt Study, MRC/Wits Rural Public Health and Heath Transitions Research Unit (Agincourt), School of Public Health, University of the Witwatersrand, Johannesburg, South Africa | |
| **Dr. Mahreen Mahmud** | Investigator | Lecturer (Assistant Professor) in Economics at the University of Exeter, Exeter, United Kingdom | |
| **Ms Zamakhanya Makhanya** | Investigator/ Trial Psychologist | Trial Psychologist DoBAt Study, MRC/Wits Rural Public Health and Heath Transitions Research Unit (Agincourt), School of Public Health, University of the Witwatersrand, Johannesburg, South Africa | |
| **Dr. Bianca Moffett** | Investigator/ Trial Manager | Trial Manager DoBAt Study, MRC/Wits Rural Public Health and Heath Transitions Research Unit (Agincourt), School of Public Health, University of the Witwatersrand, Johannesburg, South Africa | |
| **Prof. Eustasius Musenge** | Investigator/ Trial Statistician | Associate Professor of Biostatistics, School of Public Health, University of the Witwatersrand, Johannesburg, South Africa | |
| **Dr. Kate Orkin** | Investigator | Senior Research Fellow in Behavioural Economics at the Centre for the Study of African Economies, University of Oxford, Oxford, United Kingdom | |

| | | | |
|---|---|---|---|
| **Ms Julia R Pozuelo** | Investigator | Doctoral student, Department of Psychiatry, and Research Assistant in the Centre for Studies of African Economies, University of Oxford, Oxford, United Kingdom | |
| **Prof. Tholene Sodi** | Investigator | Professor, Department of Psychology at the University of Limpopo, Polokwane, South Africa | |
| **Prof. Stephen Tollman** | Investigator | Research Professor and Director, MRC/Wits Rural Public Health and Health Transitions Research Unit (Agincourt), School of Public Health, University of the Witwatersrand, Johannesburg, South Africa | |
| **Prof. Imraan Valodia** | Investigator | Pro Vice-chancellor for Climate, Sustainability and Inequality, and Director of Southern Centre for Inequalities Studies, University of the Witwatersrand, Johannesburg, South Africa | |

### Table 2. Trial Personnel and Committees

| Committee | Members and responsibilities |
|---|---|
| **Trial Management Team (TMT)** | Comprised of the co-Principal Investigators, the Trial Manager, the Trial Psychologist, and other investigators. The TMT is responsible for the day to day running of the trial and meets on a weekly basis. |
| **Risk Management Team (RMT)** | The RMT consists of the Trial Manager (a Medical Doctor), the Trial Clinical Psychologist, the Trial Registered Counsellor, and a senior Field Supervisor. Supervision and oversight are provided by Prof. Alan Stein (Child and Adolescent Psychiatrist), Prof. Kathleen Kahn (Medical Doctor), Prof. Crick Lund (Clinical Psychologist) and Prof. Tholene Sodi (Clinical Psychologist). The RMT is responsible for assessing and managing all risks amongst screened and enrolled participants. |
| **Trial Steering Committee (TSC)** | Consists of senior academic clinicians and researchers including Prof. Roz Shafran (chair of the TSC, Professor of Translational Psychology, UCL), Prof. Jonathan Roiser (Professor of Cognitive Neuroscience, UCL), Prof. Soraya Seedat (Professor of Psychiatry, Stellenbosch University), and Prof. Jonathan Levin (Professor of Biostatistics, University of Witwatersrand). The TSC oversees the scientific conduct of the study and meets on a quarterly basis throughout the trial. |
| **Independent Data and Safety Monitoring Board (DSMB)** | Comprised of Prof. Bonginkosi Chiliza (chair of the DSMB, Chief Specialist and Head of the Department of Psychiatry at the University of KwaZulu-Natal), Dr. Elizabeth George (Statistician, MRC Clinical Trials Unit at University College London), Prof. John Joska (HIV Mental Health Research Unit, Division of Neuropsychiatry, University of Cape Town), and Prof. Marguerite Schneider (Department of Psychiatry and Mental Health, University of Cape Town). The DSMB oversees the conduct and safety of the trial. Meetings will be held before recruitment begins, midway through recruitment, and at the end of the 11-week assessment period. A formal interim analysis will not be conducted. |

### Table 3. Trial registration data

| Data category | Information |
|---|---|
| **Primary registry and trial identifying number** | South African National Clinical Trials Registry (DOH-27-112020-5741); Pan African Clinical Trials Registry(PACTR202206574814636) |
| **Date of registration in primary registry** | 19th November 2020 |
| **Ethics reference** | MED20-05-011 / OxTREC 34-20 |
| **Protocol version** | v1.2 19/04/2021 |
| **Protocol amendments** | Important protocol amendments such as changes to eligibility criteria, outcomes or analysis will be reported to investigators, the TSC, and both ethics committees in writing, and relayed to study participants at the soonest availability. |
| **Funder** | MRC Newton Fund UK-South Africa Joint Initiative on Mental Health (MR/S008748/1) |
| **Sponsor** | Wits Health Consortium (Pty) Limited, University of Witwatersrand<br>31 Princess of Wales Terrace, Parktown, Johannesburg, 2193<br>011 274 9200 |
| **Trial Title** | Digital delivery of Behavioural Activation therapy to overcome depression and facilitate socio-economic transitions of adolescents in South Africa: pilot Randomised Control Trial |
| **Short Title** | DoBAt Study |
| **Countries of recruitment** | South Africa |
| **Health condition(s) or problem(s) studied** | Depression |
| **Intervention** | All participants in the intervention and control arms will be given a Samsung Galaxy A2 Core Android smartphone and receive active symptom monitoring via text messages sent to the smartphone every 2.5 weeks.<br>• The intervention arm will receive Behavioural Activation (BA) therapy via a smartphone application (the *Kuamsha* app) and supported by weekly phone calls from Trained Peer Mentors, implemented over 10 weeks.<br>• The control arm will receive a smartphone application (the *Kuchunguza* app) containing six module video clips from WildEarth-SafariLive, a locally produced wildlife series. |
| **Trial Participants** | Adolescents aged 15-19 with mild to moderately-severe depression based on scores between 5 and 19 on the 9-item Patient Health Questionnaire-Adolescent version (PHQ-A), that live in the MRC/Wits-Agincourt study area, Bushbuckridge, and from whom we obtain written informed assent and consent (including parental consent if <18 years). |
| **Study type** | Two-arm single-blind individually randomised controlled pilot trial |
| **Date of first enrolment** | 25th November 2021 |
| **Trial Status** | Enrolment of the first participant occurred on the 25th November 2021. We expect to finish recruitment during August 2022 and data collection in January 2023. |
| **Target sample size** | 200 |
| **Recruitment status** | Recruiting |
| **Primary outcomes** | 1) to determine the feasibility and acceptability of the intervention<br>2) to provide preliminary evidence on the initial efficacy (direction and magnitude) of any effects of the intervention on depressive symptoms |
| **Key secondary outcomes** | 1) to pilot a range of mental health, social-affective cognition, risky behaviours, and socioeconomic measures |

| | |
|---|---|
| | 2) to collect descriptive data on trial procedures such as recruitment, retention, data collection, randomisation and blinding to inform the development of a further larger trial. |
| **Data sharing statement** | Individual participant quantitative data that underlie the results reported in each publication arising from the trial will be made available after deidentification. Any video, audio or qualitative data will not be available. Data will be available beginning 9 months and ending 36 months following main article publication to researchers who provide a methodologically sound proposal that purposes to achieve aims in the approved proposal and /or for individual participant data meta-analysis. Data is documented and stored on the MRC/Wits-Agincourt Data Repository with a digital object identifier (doi) and can be accessed with permission and in line with with MRC/Wits-Agincourt policies and procedures. Data requestors will need to sign a data access agreement before any data can be shared. In addition, Study Protocol and Statistical Analysis Plan documents will be available. |

### Table 4. The Song Contest story

| Learning module | Description of each module | BA Learning principles |
|---|---|---|
| **Episode 1 (“Pick a team”)** | Students find out about a school Song Contest. The winning prize is a voucher to shop for a new outfit at the mall. User chooses the main character’s name and picks two teammates to join the Song Contest.<br>Main character gets anxious about the idea of performing in front of everyone else. Christine (teacher) talks about the benefits of stepping outside one’s comfort zone and offers her support.<br>At the end of the module, the user will be asked to set a goal to work on over the 10 weeks of intervention. | Absorption<br>TRAP-TRAC<br>Sleep<br>Self-confidence<br>Relapse prevention |
| **Episode 2 (“Find a cool tune”)** | First practice session with group which didn’t go well because one teammate fell asleep. Main character is frustrated and decides to ask Christine for advice. After hearing that teammate’s grandmother ill, main character showed leadership and compassion in how they handled the situation. | |
| **Episode 3 (“The Lure”)** | The team worked well together and made progress on the song. At the end of the practice, Prince (desirable character) invites the main character to go to the tavern. Main character is asked to think about the consequences of their actions but decides to go anyway. | |
| **Episode 4 (“The Fallout”)** | The next day the main character is exhausted, hungover, and unable to concentrate. They arrive late to practice session without homework done. Teammates get annoyed. Main character decides to apologise instead of avoiding the problem and their teammates are forgiving. Main character learns about the importance of sleep and goes to sleep early. | |
| **Episode 5 (“The Return”)** | Main character sleeps and feels recharged. Team is happy with the practice. Song is completed. Main character learns that they have to give a presentation if they win and asks Christine for advice. Main character learns ways to deal with being nervous and how to feel more self-confident. | |
| **Episode 6 (“The Big Day”)** | Team feels nervous before the show. They perform and win the prize. Group reflects on journey. | |

### Table 5. The Football Match story

| Learning module | Description of each module | BA Learning principles |
|---|---|---|
| **Episode 1 ("Pick a team")** | Main character is the striker for the local team. User chooses the main character's name and picks a name for the football team. User gets introduced to the football task. At the end of the module, the user will be asked to set a goal to work on over the 10 weeks of intervention. | Absorption<br>Rumination<br>TRAP-TRAC<br>Problem-solving<br>Negotiation |
| **Episode 2 ("Game Over")** | Main character misses some important shots. Shane (antagonistic character) is a discouraging main character because of their poor performance. Team loses the game. Main character leaves the match alone feeling upset and discouraged, ruminating over her performance during the match. Bird offers different, more positive perspective on performance but the main character not in a frame of mind to believe it. | |
| **Episode 3 ("Hide away")** | Main character is frustrated, misses school and gets a detention. Main character feels a bit better after friends visit her/him at her house, but apprehensive about going to school. Main character plays the football task which makes them feel energised enough to do their homework. | |
| **Episode 4 ("That's okay")** | Main character returns to school feeling more positive after a good night's sleep. Main character handles Shane well, but their mood takes a dip. Main character decides to speak with Coach Bayer after school about their performance in the match. Main character learns that mistakes can be useful opportunities to learn, instead of something to avoid. Main character practises what they've learnt and starts to feel more confident. | |
| **Episode 5 ("Keep practising")** | Main character speaks with friends before practice about what they learnt from Coach Bayer and asks for help in dealing with Shane. With their supportive presence, main character gets Shane to agree to stay away from them so that they can focus on football and winning. Main character feels confident, keeps practising, and feels ready for the re-match. | |
| **Episode 6 ("The Big Match")** | It all came together for the main character, stepping out onto the pitch with new confidence. Main character keeps calm and meets their mistakes with curiosity instead of frustration. Main character scores the winning goal. | |

**Table 6. Example mock-ups of the Kuamsha app**

| Component | Description | Example |
|---|---|---|
| *Home screen* | This is the first screen that users will see as they open the Kuamsha app. Users will have the option to play through the stories, monitor their mood, play absorbing activities to improve focus, or report on their weekly activities (see below for further details on each of these components). | 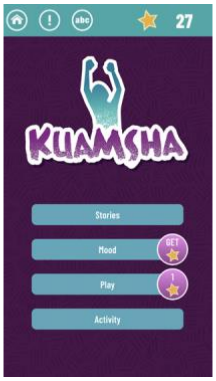  |
| *Log-in unlock code* | Kuamsha is password-protected. Users will be asked to enter a password every time they access the app. | 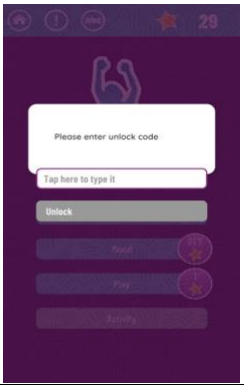  |
| *Language selector* | Users will be able to select their preferred language. All the text in the app underwent two rounds of translation and has been checked by a clinical psychologist for accuracy. | 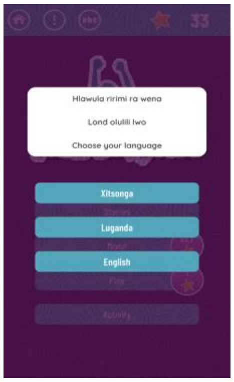  |
| *Story Selection* | The core of the game consists of a choice between two narrative stories. Each consists of 6 modules that are played in sequential order. It is possible to begin one story and then switch to the other. During gameplay points are earned for the choices made and by completing other core game elements described below. | |

| | | |
|---|---|---|
| | | 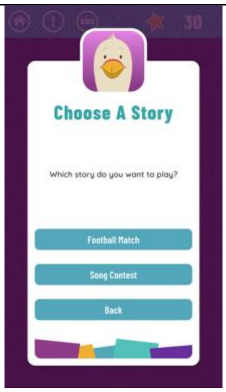 |
| *Mood monitoring* | Mood monitoring is an important part of BA and can be used as a tool for understanding how your mood is impacted and changes depending on the activities you are engaged in. Every time a user complete mood monitoring, they receive in-app points. Participants are asked to monitor their mood at various times (before and after each module, when they report a homework activity, and when they complete an absorbing activity). | 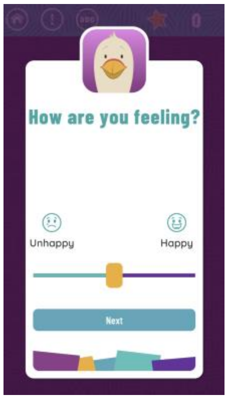 |
